# Evaluating Adoption, Impact, and Factors Driving Adoption for TREWS, a Machine Learning-Based Sepsis Alerting System

**DOI:** 10.1101/2021.07.02.21259941

**Authors:** Katharine E. Henry, Roy Adams, Cassandra Parent, Anirudh Sridharan, Lauren Johnson, David N. Hager, Sara E. Cosgrove, Andrew Markowski, Eili Y. Klein, Edward S. Chen, Maureen Henley, Sheila Miranda, Katrina Houston, Robert C. Linton, Anushree R. Ahluwalia, Albert W. Wu, Suchi Saria

## Abstract

Machine learning-based clinical decision support tools for sepsis create opportunities to identify at-risk patients and initiate treatments earlier, critical to improving sepsis outcomes. Increasing use of such systems necessitates quantifying and understanding provider adoption. Using real-time provider interactions with a sepsis early detection tool (Targeted Real-time Early Warning System) deployed at five hospitals over a two-year period (469,419 screened encounters, 9,805 (2.1%) retrospectively-identified sepsis cases), we found high sensitivity (82% of sepsis cases identified), high adoption rates (89% of alerts evaluated by a physician or advanced practice provider and 38% of evaluated alerts confirmed) and an association between use of the tool and earlier treatment of sepsis patients (1.85 (95% CI:1.66-2.00) hour reduction in median time to first antibiotics order). Further, we found that provider-related factors were strongly associated with adoption. Beyond improving system performance, efforts to improve adoption should focus on provider knowledge, experience, and perceptions of the system.

---

Clinical decision support (CDS) tools that leverage machine learning techniques are becoming more common. They have been used to facilitate early recognition of disease states, reduce diagnostic errors, and improve patient outcomes^1–4^. Of particular interest are tools that can identify at-risk patients early in the progression of a disease, allowing providers to intervene earlier and potentially improve outcomes. While traditional CDS tools use a small number of criteria to assess patient risk, tools informed by machine learning techniques use large amounts of high-dimensional historical data to learn patterns indicative of the disease of interest. They can also incorporate individual-specific features (e.g., comorbid conditions and patient history) in the algorithm. In retrospective evaluations, these systems are generally more precise and identify patients earlier in their disease trajectory^5–9^. Improved identification of disease, though, contributes little if the output is not adopted by providers^10–14^, making user adoption key to improving patient outcomes. Studies to date have shown limited success gaining widespread adoption^15–21^, with systems typically reporting users responding to 6-45% of alerts or requiring dedicated staff to review alerts and having low to moderate impact on provider practice^22–24^. However, there is limited evidence on how best to design and integrate such tools in order to improve adoption and increase impact on clinical practice.

Adoption of automated systems in non-clinical settings depends on several factors including personal characteristics and preferences of the user, characteristics of the automated system (e.g., CDS tool), and the environment in which the technology is used^25^. In clinical simulations in a ‘laboratory’ setting, where providers are shown simulated CDS recommendations for exemplar patients, studies have found that interface design^26^, provider expertise^27^, and clinical time constraints^28^ all play a role in adoption of the tool. However, in the real-world clinical setting, there are additional barriers to system adoption, including unpredictable variations in workflow, changes in personnel, and high-stakes consequences of incorrect decisions, that are difficult to replicate in simulations^29^. In this study, we sought to identify which patient, provider, and environmental factors influence adoption of a CDS tool in the real-world setting and could be modified to increase adoption of these systems.

We examined the clinical adoption of a deployed CDS tool for early detection of sepsis called the Targeted Real-time Early Warning System (TREWS). Sepsis is a life-threatening condition in which systemic infection and the host’s inflammatory response cause organ dysfunction^30^. Early recognition of sepsis is critical for successful treatment and, in particular, early administration of antibiotics is associated with decreased mortality^31–33^.

TREWS was deployed in the Johns Hopkins Health System’s two academic and three community hospitals in the Maryland/Washington D.C. area. Using electronic health record data collected between April 2018 and March 2020, we set out to answer three questions regarding clinical adoption of TREWS. First, to what degree was the tool adopted by clinicians? Second, was adoption associated with improved patient care? Answering this question is critical to understanding the success or failure of a CDS tool deployment. Third, what patient, provider, and environmental factors were associated with adoption of the tool? As additional predictive systems are deployed for facilitating proactive care, understanding the extent to which these factors impact provider adoption and dismissal of alerts for true cases can instruct how these systems should be designed.

## RESULTS

When a TREWS alert occurs on a patient, the provider (physician or advanced practice provider) has the option to open the alert and view the tool’s analysis. The bedside provider can then choose to enter an evaluation via the TREWS interface indicating whether or not they believe the patient currently has sepsis (see Methods for additional details). A primary goal of TREWS is to trigger such patient evaluations and we will use them as our primary measure of system adoption throughout.

### Study question 1: Overall adoption

During the study period, the TREWS system screened 469,419 patient encounters, 9,805 (2.1%) of which were retrospectively identified as having sepsis using the criteria listed for sepsis-related organ dysfunction in electronic health record (EHR)-based sepsis phenotyping^34,35^ (Figure 1). The system flagged 31,591 (6.7%) patient encounters for sepsis screening; average daily alert counts for each hospital are shown in Supplemental Table 1. Of the 9,805 patient encounters with sepsis, 8,033 (82%) of them were flagged by the tool. The sample characteristics for these encounters are reported in Supplemental Table 2.

**Figure 1.**
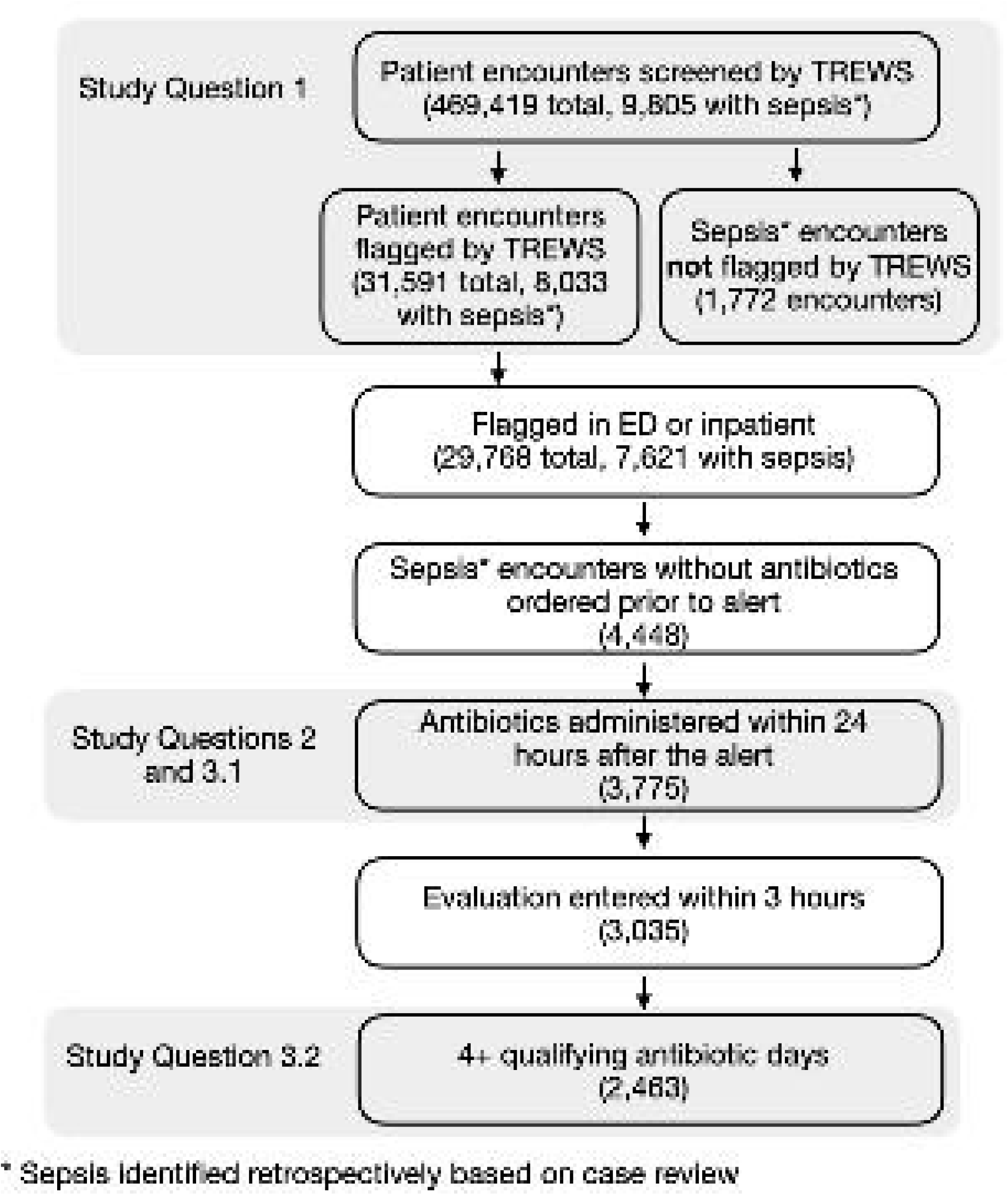
Included Study Population by Study Question: The flowchart shows the included population for each study question. Study Question 1 included 469,419 screened patients. Study Questions 2 and 3.1 included 3,775 patients with sepsis and an alert who had no antibiotic orders prior to the alert, but received antibiotics within 24 hours after the alert. Study Question 3.2 included the 2,463 of these who had an evaluation entered within 3 hours of the alert and also received a substantial antibiotics course.

A total of 1,965 providers entered at least one evaluation in TREWS during the study period. Among all patient encounters with an alert, 28,243 (89%) had a provider evaluation entered in the TREWS page, with 16,768 (53%) and 22,982 (73%) receiving evaluation within 1 and 3 hours, respectively. Alerts on patients with sepsis were evaluated at similar rates. Of the patients who had their alert evaluated at some point, 10,644 (38%) had their alert confirmed (i.e., the provider entered that the patient had sepsis at the time of evaluation). Among patients who had their alert evaluated and were retrospectively identified as having sepsis, 5,388 (71%) had their alert confirmed. The rate of confirmation was near constant across all time ranges considered. A full description of the system and its workflow is provided in the Online Methods.

### Study question 2: Impact of Adoption on Timing of Antibiotic Orders

Based on current treatment recommendations for sepsis^36^, we consider an evaluation to be ‘timely’ if it was entered within the TREWS page within 3 hours after the TREWS alert. Among retrospectively-identified sepsis patients, having a timely evaluation entered by a physician was associated with a 1.12 (95% CI 0.87 - 1.30) hour reduction in the adjusted median time from alert to first antibiotic order compared with not having a timely evaluation entered in the TREWS tool (Table 3). Further, timely alert confirmation (i.e., evaluated and confirmed within the TREWS tool within 3 hours of the TREWS alert) was associated with a 1.85 (95% CI 1.66 - 2.00) hour reduction in the adjusted median time from alert to antibiotic order compared to when alerts were not evaluated or were dismissed (Table 3).

**Table 1.** Potential Influencers of Provider Response

| Factor | Definition | Rationale for Inclusion |
| --- | --- | --- |
| **Patient Presentation Factors** |  |  |
| **Absence of Key Sepsis Symptoms** | True if no more than one of the following are met prior to the alert: lactate > 2.0 mmol/L, WBC > 12 mu/L or WBC < 4 mu/L, and temperature > 38.0°C or temperature < 36.0°C | These three criteria are commonly associated with infection and sepsis. Providers may be more willing to dismiss alerts that present without multiple of these symptoms |
| **Alternative Diagnosis** | True if any of the following diagnoses were made during the patient's stay based on the presence of ICD-10 codes: myocardial infarction, stroke, acute respiratory failure | Presence of an alternative diagnosis may increase the complexity of the diagnostic process by masking sepsis symptoms |
| **Condition at Risk for Fluid Overload** | True if any of the following chronic conditions were present based on the presence of ICD-10 codes: chronic obstructive pulmonary disorder (COPD), chronic kidney disease (CKD), congestive heart failure (CHF) | Confirming the alert is related to initiation of the sepsis bundle. Providers may dismiss the alert on patients who are at risk for fluid overload because they do not want to initiate the sepsis bundle fluid requirement |
| **Acute General Severity** | The adjustment used the raw Simplified Acute Physiology Score (SAPS) II score. For the effect estimation, this feature was true if SAPS II was above the observed median | Patients with higher SAPS II may be more complex and have other conditions that mask sepsis symptoms |
| **Chronic Complexity** | The adjustment used the raw Charlson Comorbidity Index (CCI) computed without age as a factor, since age is included as a separate factor. For the effect estimation, this feature was true if CCI excluding age, was above the observed median CCI in the population | Providers may have a higher threshold for dismissing an alert on a patient with more comorbidities because they are at a higher risk of deterioration |
| **Advanced Age** | Age > 70 years | Providers may have a higher threshold for dismissing an alert on an older patient because they are at a higher risk of deterioration |

| Environmental Factors |  |  |
| --- | --- | --- |
| **High Alert Level** | True if the total number of TREWS alerts in the past 24 hours in that unit exceeded the median for that unit and is greater than 2 alerts in the past 24 hours | Providers may have alert fatigue if there have been a lot of alerts in the past day and be less likely to respond to new alerts |
| **High Admit Volume** | True if the total number of admissions in the past 3 hours in that unit exceeded the median for that unit and the number of new admissions is greater than 2 | Providers are more busy when there are many new admissions to the unit and may be less likely to respond to alerts in a timely way |
| **Alert Occurred 7am-3pm** | True if alert occurs between 7am and 3pm | This corresponds to the morning/early afternoon hospital shift, which tends to have fewer new admissions in most units |
| **Alert Occurred 3pm-11pm** | True if alert occurs between 3pm and 11pm | This corresponds to the late afternoon/evening shift, which tends to have increased rates of new admissions and buildup of volume in the ED |
| **Alert Occurred 11pm-7am** | True if alert occurs between 11pm and 7am | This corresponds to the overnight shift, which tends to have higher total patient volume in the ED from buildup through the day, sparser provider coverage, and fewer new admissions |
| Provider Factors |  |  |
| **ED provider** | True if provider caring for the patient at the time of the alert was an ED provider | ED providers interact with patients earlier in their stay when there is more uncertainty and have a higher patient load per hour |
| **Provider Experience w/ Alert** | True if provider evaluated a previous alert within the past 30 days | Providers who are more familiar with the alert, may be more aware of the alert and be more likely to respond again |

**Table 2.** Provider Engagement with Alerts inside the TREWS Page

|  | **Alerts with provider evaluation entered**<br>**(N=31,591)** |  | **Alerts on Sepsis Cases\***<br>**(% of alerts on sepsis cases, N=8,033)** |  |
| --- | --- | --- | --- | --- |
| **Time from alert to response** | **All Alerts**<br>**(% of alerts)** | **Confirmed Alerts**<br>**(% of alerts)** | **All Alerts**<br>**(% of alerts on sepsis cases)** | **Confirmed Alerts**<br>**(% of evaluated alerts on sepsis cases)** |
| **Within 1 hour** | 16,768 (53%) | 6,184 (37%) | 4,343 (54%) | 3,162 (73%) |
| **Within 3 hours** | 22,982 (73%) | 8,587 (37%) | 5,943 (74%) | 4,311 (73%) |
| **Within 6 hours** | 25,020 (79%) | 9,337 (37%) | 6,485 (81%) | 4,680 (72%) |
| **Ever** | 28,243 (89%) | 10,644 (38%) | 7,603 (95%) | 5,388 (71%) |

\*Identified retrospectively

**Table 3.** Association of Provider Response and the Difference in Hours from Alert to Antibiotics

| **Difference between Response A vs B** | **Unadjusted median difference, hours (95% CI)** | **Adjusted median difference, hours (95% CI)** |
| --- | --- | --- |
| **Evaluation entered** w/in 3 hrs vs **No Evaluation entered** w/in 3 hrs | -1.28 (-1.50, -1.02) | -1.12 (-1.30, -0.87) |
| **Alert Confirmed** w/in 3 hrs vs **Not Confirmed** w/in 3 hrs | -1.90 (-2.02, -1.74) | -1.85 (-2.00, -1.66) |

### Study question 3.1: Factors associated with alert adoption

To further understand alert adoption, we examined which patient, provider, and environmental factors were associated with timely alert evaluation among sepsis patients. Among 3,775 sepsis patients without an antibiotic order prior to their alert and antibiotics administered within 24 hours after the alert, 3,035 had an evaluation entered within 3 hours and 740 did not. Among patient factors, advanced age (>70 years) was significantly associated with an increased likelihood of entering a timely evaluation; the remaining patient factors were not significantly associated (Table 4). Among environmental factors, alert occurrence during the 7am - 3pm shift was associated with increased likelihood of evaluating the alert. Alert occurrences between 3pm-11pm and 11pm - 3am and high admission volume were not significantly associated with adoption. However, high alert volume in the previous 24 hours was associated with decreased likelihood of evaluating the alert. Significance of the ‘High alert level’ factor did not change when a shorter time-window of 6 hours was used. Provider factors had the strongest associations with alert adoption. Emergency department providers and providers with a recent interaction with the alert had the highest likelihood of entering a timely evaluation with adjusted risk ratios of 1.22 (95% CI 1.14 -1.32) and 1.22 (95% CI 1.19 - 1.26), respectively. A complete list of factors considered is provided in Table 1.

**Table 4.** Association of factors with Evaluation of Alerts on Sepsis Patients

| **Factor** | **Unadjusted risk ratio**<br>**(95% CI)** | **Adjusted risk ratio**<br>**(95% CI)** |
| --- | --- | --- |
| **Patient Presentation Factors** |  |  |
| **Absence of Key Sepsis Symptoms** | 1.01 (0.98 - 1.04) | 0.99 (0.96 - 1.03) |
| **Alternative Diagnosis** | 0.99 (0.96 - 1.02) | 1.00 (0.97 - 1.03) |
| **Condition at Risk for Fluid Overload** | 1.02 (1.00 - 1.04) | 1.01 (0.98 - 1.04) |
| **Acute General Severity** | 0.98 (0.96 - 1.01) | 0.97 (0.94 - 1.01) |
| **Chronic Complexity** | 1.04 (1.00 - 1.08) | 1.02 (0.97 - 1.08) |
| **Advanced Age** | **1.05 (1.02 - 1.10)** | **1.06 (1.03 - 1.10)** |
| **Environmental Factors** |  |  |
| **High Alert Level** | **0.96 (0.93 - 0.99)** | **0.94 (0.91 - 0.96)** |
| **High Admit Volume** | 1.01 (0.98 - 1.05) | 0.99 (0.96 - 1.03) |
| **Alert Occurred 7am-3pm** | **1.06 (1.04 - 1.09)** | **1.03 (1.01 - 1.06)** |
| **Alert Occurred 3pm-11pm** | **0.94 (0.92 - 0.97)** | 0.98 (0.95 - 1.00) |
| **Alert Occurred 11pm-7am** | 1.00 (0.95 - 1.03) | 1.01 (0.97 - 1.04) |
| **Provider Factors** |  |  |
| **ED provider** | **1.35 (1.24 - 1.49)** | **1.22 (1.14 - 1.32)** |
| **Provider Experience w/ alert** | **1.25 (1.21 - 1.29)** | **1.22 (1.19 - 1.26)** |

### Study question 3.2: Factors associated with incorrect alert dismissal

Even when a provider responds to an alert, sepsis may not be recognized. As such, we examined which patient, provider, and environmental factors were associated with alert dismissal on sepsis patients who received a substantial antibiotic course (4+ continuous days of an antibiotics or antibiotics until death or transfer to an acute care facility). See Online Methods for details. Among the alerts on included patients with sepsis (N=7,621), 2,463 received a timely evaluation and met the additional 4+ antibiotic day restriction (1,751 confirmed alerts and 712 dismissed alerts). Among patient factors, the absence of key sepsis symptoms and younger age were significantly associated with an increase in the likelihood of dismissing the alert (Table 5). High acute general severity was also associated with an increase in the likelihood of dismissing the alert. Other patient factors were not significantly associated with alert dismissal. Among provider factors, working in the ED and recent interactions with alerts were both associated with decreased likelihood of dismissal and, among environmental factors, alerts occurring during the evening or overnight shifts (3pm - 11pm or 11pm - 7am) were more likely to be dismissed.

**Table 5.** Association of factors with Dismissal of Evaluated Alerts on Sepsis Patients

| Factor | Unadjusted risk ratio<br>(95% CI) | Adjusted risk ratio<br>(95% CI) |
| --- | --- | --- |
| **Patient Presentation Factors** |  |  |
| **Absence of Key Sepsis Symptoms** | 1.01 (0.86 - 1.19) | **1.28 (1.06 - 1.45)** |
| **Alternative Diagnosis** | **1.27 (1.14 - 1.42)** | 1.11 (0.97 - 1.32) |
| **Condition at Risk for Fluid Overload** | 1.10 (0.97 - 1.21) | 1.08 (0.97 - 1.22) |
| **Acute General Severity** | **1.39 (1.23 - 1.56)** | **1.46 (1.28 - 1.66)** |
| **Chronic Complexity** | **0.87 (0.76 - 0.98)** | 0.90 (0.75 - 1.05) |
| **Advanced Age** | **0.74 (0.65 - 0.81)** | **0.69 (0.60 - 0.75)** |
| **Environmental Factors** |  |  |
| **High Alert Level** | 0.91 (0.80 - 1.01) | 1.01 (0.90 - 1.13) |
| **High Admit Volume** | **0.83 (0.73 - 0.94)** | 0.98 (0.86 - 1.12) |
| **Alert Occurred 7am-3pm** | **0.87 (0.74 - 0.99)** | 1.12 (0.99 - 1.28) |
| **Alert Occurred 3pm-11pm** | 1.04 (0.92 - 1.16) | **1.20 (1.09 - 1.33)** |
| **Alert Occurred 11pm-7am** | **1.15 (1.03 - 1.29)** | **1.19 (1.07 - 1.36)** |
| **Provider Factors** |  |  |
| **ED provider** | **0.39 (0.34 - 0.43)** | **0.47 (0.40 - 0.54)** |
| **Provider Experience w/ alert** | **0.58 (0.48 - 0.64)** | **0.66 (0.56 - 0.73)** |

## DISCUSSION

In this study, we characterized the adoption and clinical impact of TREWS, a machine learning-based clinical decision support system for sepsis, and evaluated the extent to which patient presentation, environmental, and provider-related factors were associated with provider reaction to the alert. TREWS was adopted at a high rate, with providers entering evaluations for 89% of alerts (73% of alerts within 3 hours), with 37-38% of those patients confirmed by the provider as having sepsis. Timely confirmation of alerts was associated with a shorter time from alert to first antibiotic order (−1.85 hours (CI -2.00, -1.66)) among patients with sepsis. Studies have found that every hour delay in sepsis treatment is associated with significant increase in mortality^32,33,37^; the observed reduction in time to antibiotics suggests that use of the tool as intended can lead to faster treatment among sepsis patients and improved clinical outcomes. Analysis of the associations between patient presentation, alert environment, and provider characteristics and real-time provider response to alerts, showed that provider characteristics had the strongest association with the decision to evaluate the alert. However, among alerts with timely adoption, certain patient, provider, and environmental factors were significantly associated with a provider’s confirmation of the alert.

In sepsis, based on promising retrospective validation, a growing number of tools have been deployed prospectively^20,38–43^. A subset of these have shown process impact^38,40,41^ but had to rely on dedicated staff to manage the high alert volumes and false alarm rates. Employing dedicated staff can ensure adoption, but poses challenges for scaling CDS to monitoring multiple conditions. Instead, deploying reliable CDS with low alert volumes that are designed to integrate into the clinical workflow and encourage adoption can enable bedside implementation that improves responsiveness and value of the alerts, while both reducing alert burden and the cost of additional staff.

The high overall rate of provider response to the TREWS alert observed in this study (a provider entered an evaluation in response to 89% of alerts) is promising given the documented challenges to gaining adoption of such systems^11,20,21,44–46^. Alert burden and the perceived accuracy of a CDS tool both play major roles in tool adoption and trust^15,16,29^ and tuning a system to achieve the highest possible performance remains critical to a successful deployment. One reason for the high observed adoption of TREWS may be the high predictive performance and low alert burden of TREWS relative to comparable deployed systems. Even with a sensitivity of 82%, precision was high with 1 in 3 evaluated alerts confirmed by a provider to have sepsis. Past deployed systems have reported significantly lower predictive performance on similar hospital populations^20,42,45^. For example, one of the most widely deployed sepsis early warning systems had a sensitivity of only 33% and a precision of 2.4% (1 in 46 alerts within 24 hours of sepsis onset)^42^. Additionally, ease of use and integration into the workflow have been noted as important factors influencing adoption^26,28,47,48^. Availability of TREWS within a provider’s EHR workflow and the inclusion of alert context to avoid “black box” presentation may have also improved overall adoption of the tool.

Provider characteristics had the strongest association with the likelihood of evaluating a TREWS alert. To a lesser extent, environmental factors like time of day were also associated with the likelihood of evaluation, and we did not find an association between patient presentation and alert evaluation. Providers who work in the ED or who had previously interacted with the tool and entered an evaluation, were most likely to evaluate a new alert. There are several possible reasons for these results. Some providers may be more willing to adopt new CDS tools than others; this tendency is sometimes referred to as “dispositional trust”^49,50^. Additionally, increased familiarity with the system may add to its perceived ease of use or accuracy. Since most first alerts occurred in the ED, those providers may naturally get more exposure to TREWS and be more familiar with the system. This is an example of “learned trust”^50^. Alternatively, the environment of the ED differs from inpatient units in several relevant ways. First, the higher degree of uncertainty around patients and emphasis on protocolized sepsis treatment (the Centers for Medicare and Medicaid Services’ ‘SEP-1 sepsis bundle’) in the ED may increase provider willingness to utilize the alert. Second, the workflow within the ED requires that providers make more consistent contact with the EHR system, increasing the likelihood that providers see and thus respond to alerts. Both of these are examples of “situational trust”^50^. Creating opportunities to interact with and practice using TREWS in a simulated setting or adapting the alert policy and interface design for different types of providers could help increase familiarity and increase adoption. Finally, providers who work in the ED will generally see more cases and have less information on each individual case. Previous work has shown that an increased workload leads to increased reliance on automated tools^50^ and thus, increased adoption in the ED may be expected and appropriate to the treatment context.

The lack of an association between patient factors and the likelihood of a provider entering a timely evaluation could be viewed as promising. It suggests that providers are willing to engage with the system even in cases that do not display an obvious presentation of sepsis. However, patient presentation *was* associated with alert dismissal on sepsis patients. We found that alerts occurring on sepsis patients who did not have certain key sepsis symptoms or with higher acute complexity at the time of the alert, were more likely to have their alert dismissed. It makes sense that alerts are more likely to be confirmed when there is clear support for the diagnosis and a lack of alternate explanation. However, this may pose a problem in cases where patients have less typical presentations of sepsis or where the alert occurs in advance of those symptoms developing. Further, if TREWS is perceived as less accurate in cases with high general acute severity, adoption may be lower in these cases as well. Education to increase awareness about alternate presentations of sepsis or situations where patient complexity may mask developing sepsis symptoms, may help improve provider trust in the system and understanding that alerts are delivering valuable information.

Among environmental factors, alert dismissal was most strongly associated with time of day, with alerts occurring during the 3pm-11pm or 11pm-7am shifts more likely to be dismissed, even after accounting for patient presentation. This may reflect an association between time of day and unit volume. In the ED, total patient volume and workload generally increases throughout the day, peaking in the evening, but remaining high even through 2-3am. Greater workload during the later shifts could contribute to a perception that dismissing the alert is faster than evaluating and completing related documentation on the TREWS page. Increasing awareness about the benefits of timely evaluation and creating supplemental support teams during peak hours, could improve uptake of ML-based CDS systems during these times.

During the study period, 1,965 providers responded to TREWS alerts. To our knowledge, this is the first prospective study of a deployed machine learning-based bedside clinical tool that quantitatively studies and achieves high provider adoption. Adoption of CDS has been studied across a wide range of clinical applications^24,51^. These studies generally report low to moderate adoption, with clinicians responding to anywhere from 6 to 45% of alerts depending on the clinical task, interface design, and workflow integration^22–24,52^. As expected, the lack of adoption typically translates to low to moderate impact on clinical processes^24,51^. In order to better understand factors impacting the adoption of CDS, previous studies of deployed systems have used post-hoc surveys or interviews to gauge provider impressions^28,44,53^; however, provider impressions can differ from their actual use^4,18,27^. A common alternative is to study real-time provider response using a clinical simulation^26,27^. This allows researchers to study the use of the tool under varying design choices. However, the simulated environment is unable to capture the full complexity of high-risk decision-making in a live-care delivery setting like the hospital environment, where there are many competing time and attention demands^4,15,16,29^. As a result, the factors found to influence response may not generalize to the practice setting. By using real-time interactions with a deployed clinical support system, we were able to assess the extent to which different factors influenced real-time decision making and treatment at a large scale and inform future system design.

This study had several limitations. First, there is a lack of consensus on how best to identify sepsis retrospectively. In order to maximize the reliability of sepsis labels, we identified sepsis cases using an EHR-based sepsis phenotype which accounts for confounding comorbidities and has shown increased sensitivity and precision compared to alternatives^34,35^. However, we cannot completely exclude the possibility that some patients had non-infectious syndromes mimicking sepsis. We also added requirements for a significant antibiotic course when identifying incorrect dismissals of alerts on identified sepsis cases. Second, we relied on International Classification of Diseases-10 (ICD-10) codes to identify the presence of chronic conditions and alternative diagnoses. While common in large retrospective studies, this may introduce some bias from coding practices. Third, all hospitals in this study were part of the same health system. However, the study includes a large cohort representing a diverse patient population from both academic and community hospitals. Fourth, this study focuses on quantitative evaluation of provider interactions that were recorded within the tool itself and does not capture any sepsis-related discussions or actions that occurred outside the tool. Finally, this study assesses the extent to which each of the factors affects adoption in the context of the deployed TREWS system, which has specific performance characteristics, interface presentation, and policy decisions about how to integrate alerts into clinical workflow. The relative importance of different factors may vary depending on the performance characteristics of the system. Additionally, increased deployment of data driven CDS systems may change provider attitudes in the future.

An additional study using qualitative human factors tools to study provider perception is underway. While we incorporated information about provider type and experience in the tool, we were unable to access additional information about provider background and attitudes towards CDS. Further study is needed to understand how these additional characteristics may affect overall adoption and the potential for alert adoption to lead to over-reliance on the alerts (e.g., over-prescription of antibiotics in response to sepsis alerts). Quantifying over-prescription resulting from a system is important for understanding the potential harms of a system^54^; however, we currently lack metrics to assess over-prescription and leave this to future work.

## CONCLUSION

Using real-time interactions with a ML-based sepsis support system, we characterized the adoption and clinical impact of the tool and identified key factors related to failure to use the tool. Overall, TREWS showed high provider adoption and significant impact on a key clinical process metric of reducing time to antibiotics for sepsis patients. Analysis of factors driving adoption showed provider-related factors, such as past experience with the system and working in the emergency department where providers had increased exposure to the system had the strongest association with willingness to evaluate alerts. While patient presentation factors like patient severity and absence of key sepsis symptoms were not significantly associated with the likelihood of evaluation, they did impact the likelihood of dismissing the alert. Education to increase awareness of how patient presentation may encourage providers to accept recommendations on sepsis cases with less common presentation. In addition to improving model performance, future ML-based systems should focus on the provider in their design choices to encourage adoption and realize the potential benefit of these systems.

## METHODS

This study was approved by the Johns Hopkins University internal review board (IRB No. 00252594) and a waiver of consent was obtained.

### Targeted Real-time Early Warning System (TREWS)

#### Description of the TREWS Model

TREWS is a machine learning-based early warning system and decision support tool that was trained using historical electronic health record (EHR) data to recognize sepsis early in its progression. The system uses routinely-collected laboratory measurements, vital signs, notes, medication history (excludes antibiotics), procedure history, and clinical history from the EHR to generate patient-specific sepsis risk score and alerts^6,55^. To improve alert performance, the system uses several machine learning-based techniques for tuning to patient context^56^, handling missing data^57^, suppressing untrustworthy alerts^58^ and improving reliability and transportability^59,60^.

#### Deployment Process

Prior to deployment at a new hospital, the alert threshold was set to achieve an 80% sensitivity at that hospital based on applying the model to historical data from that hospital. The same model parameters were used at each site. The deployment at each hospital was done in three steps. First, a team of educators including clinicians from the site and members of the tool development team, met with clinicians to explain the tools functionality, identify clinical champions, and to verify the process for clinical workflow integration. During this period, the alert was active in the background and the technical team monitored the alert volume across different subpopulations in the hospital. Second, deployment was piloted in order to verify the integration of the system at each site with a subset of the users. Finally, the alert was activated in all ED and inpatient units and the deployment entered a maintenance stage. Throughout the deployment process and maintenance period, the technical and clinical teams monitored alert performance and provider use in different units through weekly emails summarizing alert interactions, performance, and alert volume. Alerts included in the analyses in this study occurred after activating the alert in all units.

#### TREWS Workflow

To minimize workflow interruptions and alert fatigue, TREWS uses a passive approach to signal new alerts. Instead of triggering a pop-up box or a pager message, the system flags patients visually within the EHR, but does not actively interrupt the provider or require an immediate response before allowing the provider to continue using the EHR. Once the alert appears (e.g., as an icon within the clinician’s patient list), a provider (physician or advanced practice provider) can click an icon to address the alert leading to a real-time workflow within the patient chart. From within this workflow, the provider can view summary data gathered by TREWS including factors leading to why the alert was generated, probability measures indicating likelihood of mortality and sepsis, and the status of sepsis-related treatments. Providers are asked to enter an evaluation of whether or not they believe the patient currently has sepsis; however, the response is not mandated. A nurse can also pre-screen an alert and escalate it to a provider if there are indications of new or worsening infection or altered mental status. There are three levels of alert response in the TREWS workflow. First is the choice to evaluate the patient following an alert and to enter an evaluation within the tool. Second is a judgement of whether the provider believes the alert is correct and if the patient does, indeed, have sepsis. Third is treatment of the patient, which will necessarily reflect their belief in the alert and diagnosis of the patient.

### Study Population

The study population included all adults who presented to the emergency department (ED) or were admitted to a medical or surgical unit at any of five hospitals (three community and two academic hospitals) in the Maryland-DC area that either 1) had a prospective TREWS alert or 2) were retrospectively identified as having sepsis based on specified criteria. The included hospitals and date ranges were: Howard County General Hospital (April 1, 2018 - March 31, 2020), Suburban Hospital (October 1, 2018 - March 31, 2020), Bayview Medical Center (February 1, 2019 - March 31, 2020), Johns Hopkins Hospital (April 1, 2019 - March 31, 2020), and Sibley Memorial Hospital (May 1, 2019 - March 31, 2020). The start date at each hospital was based on the timing of the staggered deployment across the five sites. We treated each time a patient presented to the ED or was admitted as a unique patient encounter and included each encounter separately. Population characteristics and overall adoption rates (Study question 1) were estimated using all patient encounters with an alert or sepsis diagnosis during this period. Based on a refinement of the criteria used in the third sepsis consensus definition (Sepsis-3)^30^ and the CDC Adult Sepsis Event Toolkit sepsis criteria^61^, sepsis cases were retrospectively identified using EHR-based sepsis phenotyping, which identifies patients with sepsis based on clinical symptoms and orders indicating suspected infection and related acute organ dysfunction within 48 hours of each other, while also adjusting for the effects of confounding comorbidities on these criteria^34,35^.

When evaluating the association between adoption and clinical care and between various factors and adoption or confirmation (Study questions 2, 3.1, and 3.2) we only included sepsis patients who received an alert in the ED or an inpatient unit (e.g., patients who had their alert before being assigned a bed in the ED were excluded) and had not received an antibiotic order at the time of their alert. This was done to restrict the analysis to cases where there was opportunity for the alert to impact care decisions. To further ensure that the antibiotic order was related to the alert, we only included patients who received antibiotics within 24 hours after the alert.

### Key Definitions

Since the goal of the early warning system is to trigger an evaluation of the patient for sepsis, our primary measure of adoption was whether or not the provider entered a patient evaluation (either confirmed as having sepsis or dismissed as not having sepsis) within the tool following the alert. We considered an evaluation “timely” if it was entered within three hours after the alert. The three hour window was chosen to match the treatment window recommended by the Centers for Medicare and Medicaid Services (CMS) sepsis core measure (SEP-1) and the Surviving Sepsis Campaign guidelines^36,62,63^. Since providers were not required to respond to alerts within the TREWS interface, this definition may not capture all patient evaluations resulting from the TREWS alerts (e.g., a provider may see the alert in the EHR and choose to document and initiate sepsis treatment without documenting it in the tool interface). However, since the vast majority of alerts had an evaluation entered in our study (see the Results section for more details), we consider this to be a strong proxy measure.

### Study Questions and Approach

#### Study question 1: Assessing the degree of adoption

To understand the alerting behavior of TREWS, we first report the number of patients screened by the system and the percentage of encounters with sepsis and/or with an alert. We then report the number and percentage of alerts with evaluations entered within 1, 3, and 6 hours after the alert or ever evaluated to understand the adoption of TREWS. Additionally, among alerts with an evaluation entered, we report the percentage that were confirmed prospectively as having sepsis. We report these numbers for all patients with an alert and patients who were retrospectively identified as having sepsis based on automated case identification, as described above^34^.

#### Study question 2: Assessing the association between adoption and patient care

To assess the association between tool adoption and patient care, we examine the extent to which using the TREWS page to record an evaluation for sepsis within three hours after the alert was associated with differences in the timing of first antibiotic administration, a key element of sepsis treatment^32,33,64^. We estimated the change in median hours from alert to antibiotics using a quantile regression. We estimated both the unadjusted median and an adjusted median that accounts for the adjustment covariates listed below (Table 1). We repeated this analysis to compare the adjusted risk difference between confirmed alerts vs those where the alert was either not evaluated within three hours or was dismissed. All models and statistics were computed using Python (version 3.7.6). The quantile regression was computed using the StatsModels Python package (version v0.12.2)^65^. We used bootstrap resampling with 3,775 bootstrap samples and 100 iterations to compute 95% confidence intervals (CI).

#### Study question 3.1: Assessing which patient, provider, and environmental factors are associated with alert adoption

To assess the impact on alert response of patient presentation and history, unit and alert environment factors, and provider characteristics on alert response, we measured the association between these factors and whether or not a patient evaluation was entered within three hours after the alert. Specific factors that might affect alert response were identified based on clinical feedback from emergency department, intensive care unit, and general ward providers actively using the tool and who had experience managing sepsis patients (Table 1). Patient factors included age, chronic complexity as measured by age and the Charlson Comorbidity Index (CCI)^66^, and acute severity as measured by the Simplified Acute Physiology Score (SAPS) II (Table 1). We also accounted for presence of sepsis-related symptoms, an alternative diagnosis that may complicate sepsis diagnosis, and the presence of chronic condition(s), such as COPD, CHF, or CKD, that may make a provider hesitant to follow the sepsis bundle guidelines for giving high-volume fluids. We characterized environmental factors based on the shift during which the alert occurs, the TREWS alert burden in the unit, computed as the number of alerts that occurred in that unit in the past 24 hours, and the admit volume computed as the number of new patients admitted to that unit in the past 3 hours. Provider factors included prior experience with TREWS and location of care provision (emergency department vs inpatient). Due to the low number of inpatient alerts, we were unable to further divide inpatient providers into medical and surgical providers.

For each factor, we estimated the adjusted and unadjusted risk ratio using a logistic regression model^67^. The adjusted model included all listed patient presentation, environmental, and provider factors (Table 1). We used nonparametric bootstrap resampling with 100 bootstrap replicates to estimate percentile-based 95% CIs. All statistical analyses were done in Python (version 3.7.6).

#### Study question 3.2: Assessing which patient, provider, and environmental factors are associated with alert dismissal on sepsis patients

To assess which patient, provider, and environmental presentation factors are associated with a provider’s decision to dismiss an alert on a patient later identified as having sepsis, we estimated the association between these factors and the evaluation entered for sepsis patients with an alert entered within three hours. As before, we excluded all patients who received an antibiotic order prior to the alert and also excluded patients with no evaluation within three hours. Since this question examines factors related to incorrect dismissal, we chose to use a more conservative inclusion criteria and restricted the study population to only the sepsis patients who received a substantial antibiotic course, namely four consecutive days of antibiotics or antibiotics up until the time of in-hospital death, discharge to hospice, or transfer to another acute care facility. Antibiotics included any of the antibiotics listed in the CMS SEP-1 core measure. We refer to this criteria going forward as having “4+ qualifying antibiotic days”. We assessed all previously described patient factors and adjusted for all patient, provider, and environmental factors. Adjusted risk ratios and associated confidence intervals were estimated as above.

## Data Availability

The data is not available for public use.

## Acknowledgements

The authors would like to thank Hossein Soleimani, Yanif Ahmad, Andong Zhang, Maxwell Yeo, and Yan Karklin whose work significantly contributed to early iterations of the development of the deployed system. Further, we wish to thank Renee Demski, Karen D’Souza, Allen Kachalia, Allen Chen, and clinical and quality stakeholders who contributed to tool deployment, education, and championing the work. This work was supported by funding from the Gordon and Betty Moore Foundation (Grant no. 3186.01), the National Science Foundation (Grant no. 1840088), and the Sloan Foundation. This information or content and conclusions are those of the authors and should not be construed as the official position or policy of, nor should any endorsements be inferred by the NSF the U.S. Government.

## Author Contributions

KEH, RA, CP, ESC, AWW, and SS contributed to the initial study design and preliminary analysis plan. KEH, AS, RCL, LJ, MH, SM, DNH, AWW, and SS contributed to the system deployment. KEH, RA, CP, EYK, SEC, ARC, ESC, DNH, AWW contributed to the review and analysis of the results. All authors contributed to the final preparation of the manuscript.

## Competing Interests Statement

Under a license agreement between Bayesian Health and the Johns Hopkins University, Dr. Henry, Dr. Saria, and Johns Hopkins University are entitled to revenue distributions. Additionally, the University owns equity in Bayesian Health. This arrangement has been reviewed and approved by the Johns Hopkins University in accordance with its conflict of interest policies. Dr. Saria also has grants from Gordon and Betty Moore Foundation, the National Science Foundation, the National Institutes of Health, Defense Advanced Research Projects Agency, the Food and Drug Administration, and the American Heart Association; she is a founder of and holds equity in Bayesian Health; she is the scientific advisory board member for PatientPing; and she has received honoraria for talks from a number of biotechnology, research, and health-tech companies. This arrangement has been reviewed and approved by the Johns Hopkins University in accordance with its conflict of interest policies. Dr. Hager discloses salary support and funding to his institution from the Marcus Foundation for the conduct of the Vitamin C, Thiamine, and Steroids in Sepsis Trial. Dr. Cosgrove consulting fees from Basilea for work on an infection adjudication committee for a *S. aureus* bacteremia trial. The other authors declare no disclosures of conflicts of interest.

## Funding

The authors gratefully acknowledge the following sources of funding: the Gordon and Betty Moore Foundation (award #3926), the National Science Foundation Future of Work at the Human-technology Frontier (award #1840088), and the Alfred P. Sloan Foundation research fellowship (2018).

