## Supplemental Materials for "Evaluating Adoption, Impact, and Factors Driving Adoption for TREWS, a Machine Learning-Based Sepsis Alerting System"

Supplemental Table 1. TREWS Alert Volume Per Day During the Study Period including re-alerts and alerts flagging patients who are candidates for escalation.

| Hospital | Number of beds | Average alerts per day | Average ED alerts per day | Average IP alerts per day |
|---|---|---|---|---|
| HCGH | 243 | 18.8 | 9.9 | 9.0 |
| SH | 228 | 15.9 | 7.7 | 8.2 |
| BMC | 463 | 21.1 | 11.3 | 9.8 |
| JHH | 1162 | 38.2 | 12.1 | 26.2 |
| SMH | 288 | 8.9 | 4.1 | 4.8 |

Supplemental Table 2. Population Characteristics

| | All Patients with an Alert (N=31,591) | Patients Included in Factor Analysis* (N=3,775) |
|---|---|---|
| Median age, years (IQR) | 66 (53-78) | 69 (56-80) |
| Male (%) | 16,336 (52%) | 1,993 (53%) |
| Median SAPS II (IQR) | 37 (27-49) | 46 (35-62) |
| Median CCI (IQR) | 5 (2-7) | 5 (3-8) |
| CHF (%) | 5,624 (20%) | 714 (20%) |
| CKD (%) | 8,859 (32%) | 1,127 (32%) |
| ESRD (%) | 2,039 (7%) | 269 (8%) |
| COPD (%) | 6,550 (24%) | 806 (23%) |
| Died in-hospital | 2,276 (7%) | 659 (17%) |
| Median Hospital LOS, days (IQR) | 4 (2-8) | 6 (4-12) |
| Ever admitted to the ICU (%) | 8,781 (28%) | 1,858 (49%) |
| Discharged from ED** | 3,053 (10%) | 52 (1%) |
| Trauma admission | 306 (1%) | 10 (0%) |
| Sepsis Case*** | 8,033 (25%) | 3,775 (100%) |
| 4+ Qualifying Antibiotic Days**** | 11,396 (36%) | 3,060 (81%) |
| Number of distinct providers who entered an evaluation on an alert | 1,965 | 627 |

IQR: interquartile range, SAPS: Simplified Acute Physiology Score, CCI: Charlson Comorbidity Index, CHF: congestive heart failure, CKD: chronic kidney disease, ESRD: end-stage renal disease, COPD: chronic obstructive pulmonary disorder, LOS: length of stay, ICU: intensive care unit, ED: emergency department

* Includes all sepsis patients with an alert who did not have an antibiotic order prior to the alert and who received antibiotics within 24 hours after the alert

** Includes patients who died in the ED or who were transferred to hospice or another acute care facility

*** Sepsis identified retrospectively based on clinical presentation (see Methods)

**** 4+ continuous days of antibiotics or antibiotics given up until day of discharge to hospice, another acute care facility, or in-hospital death
